# A single-nucleus transcriptome-wide association study implicates novel genes in depression pathogenesis

**DOI:** 10.1101/2023.03.27.23286844

**Authors:** Lu Zeng, Masashi Fujita, Zongmei Gao, Charles C. White, Gilad S. Green, Naomi Habib, Vilas Menon, David A. Bennett, Patricia A. Boyle, Hans-Ulrich Klein, Philip L. De Jager

**Affiliations:** Center for Translational and Computational Neuroimmunology, Department of Neurology, Columbia University Irving Medical Center, New York, NY, USA; Edmond & Lily Safra Center for Brain Sciences, The Hebrew University of Jerusalem, Jerusalem, Israel; Rush Alzheimer Disease Center, Rush University Medical Center, Chicago, Illinois, USA; Department of Neurological Sciences, Rush University Medical Center, Chicago, Illinois; Department of Psychiatry and Behavioral Sciences, Rush University Medical Center, Chicago, Illinois

## Abstract

**Background:** Depression is a common psychiatric illness and global public health problem. However, our limited understanding of the biological basis of depression has hindered the development of novel treatments and interventions.

**Methods:** To identify new candidate genes for therapeutic development, we examined single-nucleus RNA sequencing (snucRNAseq) data from the dorsolateral prefrontal cortex (N=424) in relation to ante-mortem depressive symptoms. To complement these direct analyses, we also used genome-wide association study (GWAS) results for depression (N=500,199) along with genetic tools for inferring the expression of 22,159 genes in 7 cell types and 55 cell subtypes to perform transcriptome-wide association studies (TWAS) of depression followed by Mendelian randomization (MR).

**Results:** Our single-nucleus TWAS analysis identified 71 causal genes in depression that have a role in specific neocortical cell subtypes; 59 of 71 genes were novel compared to previous studies. Depression TWAS genes showed a cell type specific pattern, with the greatest enrichment being in both excitatory and inhibitory neurons as well as astrocytes. Gene expression in different neuron subtypes have different directions of effect on depression risk. Compared to lower genetically correlated traits (e.g. body mass index) with depression, higher correlated traits (e.g., neuroticism) have more common TWAS genes with depression. In parallel, we performed differential gene expression analysis in relation to depression in 55 cortical cell subtypes, and we found that genes such as *ANKRD36*, *MADD*, *TAOK3*, *SCAI* and *CHUK* are associated with depression in specific cell subtypes.

**Conclusions:** These two sets of analyses illustrate the utility of large snucRNAseq data to uncover both genes whose expression is altered in specific cell subtypes in the context of depression and to enhance the interpretation of well-powered GWAS so that we can prioritize specific susceptibility genes for further analysis and therapeutic development.

## Introduction

Depression is a common psychiatric illness and is the third leading cause of years lived with disability worldwide (1, 2). Alleviating the burden of this costly disease is an important priority; however, our limited understanding of the biological basis of depression has hindered the development of novel treatments and interventions.

Despite the impressive success of the genome-wide association studies (GWAS), there is a substantial gap between the susceptibility variants discovered and understanding how those susceptibility loci contribute to disease onset. Most of the GWAS signals map to non-coding regions and, often, a locus’ local linkage disequilibrium (LD) structure does not permit firm conclusions about the identity of the causal variant and its functional effects, which poses a challenge for the identification of risk genes (3) and cell types in which these risk variants may alter gene expression. One approach for addressing this variant-to-function challenge is to use expression quantitative trait loci (eQTL) mapping to characterize the impact of disease-associated regulatory variants on the expression of nearby genes (4).

Given the complexity of psychiatric disorders such as depression, disentangling the role of each cell type in the brain is important and requires studies performed at single-cell resolution. In this study, to identify potential causal depression genes in brain, we have leveraged a large set of data on individual nuclei extracted from the dorsolateral prefrontal cortex (DLPFC), an important hub in mood circuits, of 424 older individuals from a collection of prospectively collected brain autopsies of the Religious Orders Study and Rush Memory and Aging Project (ROSMAP) cohort. We integrated this gene expression data set and results of depression GWAS analyses (5), as implemented in function summary-based imputation (FUSION) (6), Summary-data-based Mendelian Randomization (SMR) (7) and colocalization analysis (COLOC) (8) in the 7 major cell types of the DLPFC. In this study, FUSION identifies genes whose cis-regulated gene expression is associated with depression, and SMR tests whether these genes mediate the association between genetic variants and depression, COLOC assesses how replicable of FUSION analysis. FUSION is a suite of tools for performing transcriptome-wide association studies (TWAS), it builds predictive models of the genetic component of a functional phenotype and predicts and tests each component for association with disease using GWAS summary statistics. COLOC is used to perform genetic colocalization analysis of two potentially related phenotypes, to ask whether they share common genetic causal variant(s) in a give region. TWAS and COLOC had similar power under the scenario with a single typed causal variant, but TWAS had superior performance when the causal variant was untyped or in the presence of allelic heterogeneity (6). SMR integrates summary-level data from GWAS with data from eQTL studies to identify genes whose expression levels are associated with a complex trait because of pleiotropy.

Furthermore, we also performed differential gene expression analysis within 55 cortical cell subtypes that were present in >=100 participants, discovering additional depression-associated genes expressed only in a subtype of cells, highlighting the importance of cellular context for uncovering many variants which influence gene expression (**Figure 1**). Our analyses illustrate the utility of large snucRNAseq data to uncover both genes whose expression is altered in specific cell subtypes in the context of depression and to enhance the interpretation of well-powered GWAS so that we can prioritize specific susceptibility genes for further analysis.

**Figure 1.**
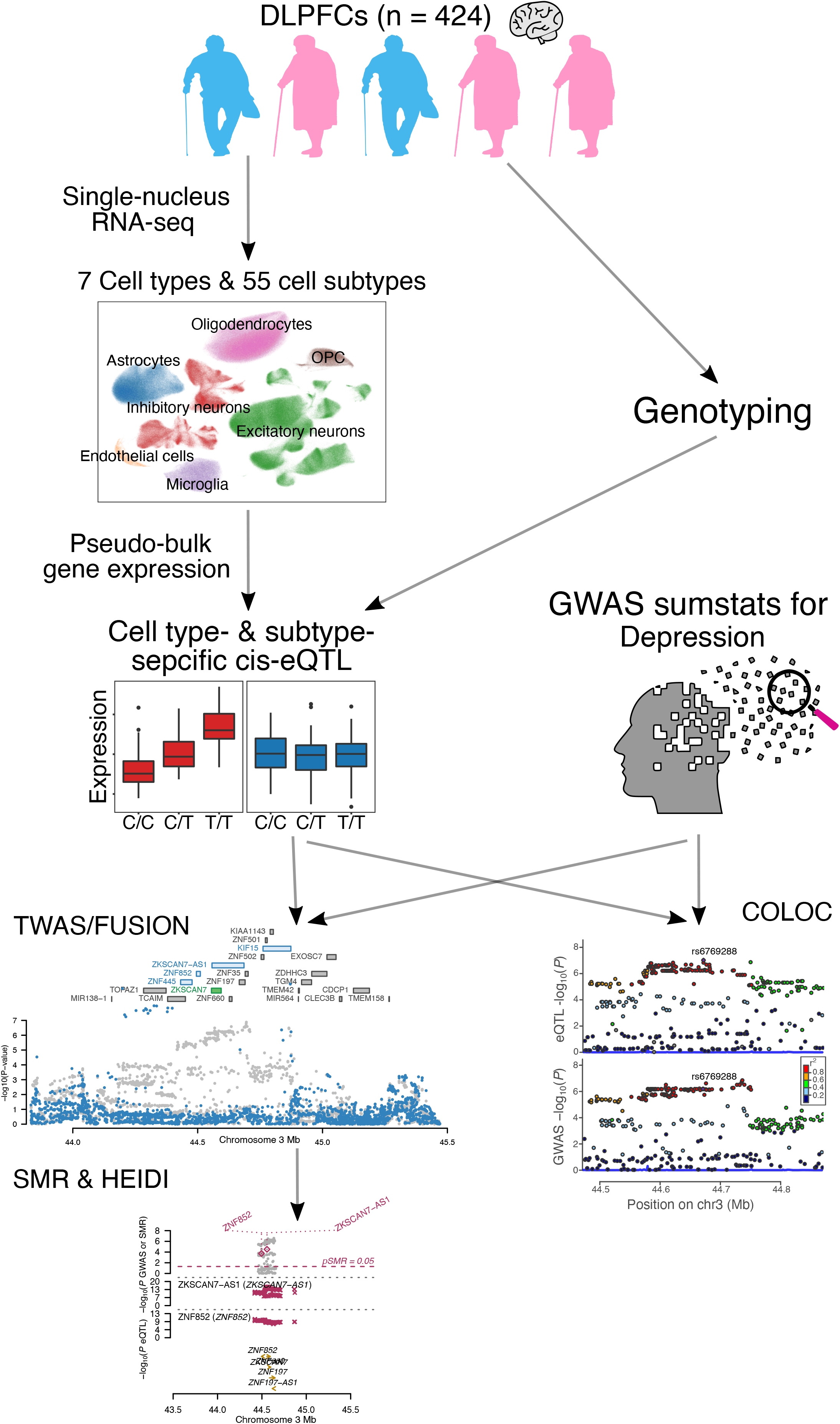
Study overview. We leveraged a large set of data derived from individual nuclei extracted from the DLPFCs of 424 genotyped older individuals. We integrated gene expression data and results of depression GWAS analyses to perform transcriptome-wide association analysis (TWAS), mendelian randomization (MR) and colocalization analysis (COLOC) in the 7 major cell types and 55 cell subtypes of the DPLFPC. Differential gene expression analysis was applied to discover additional depression associated genes expressed only in a subtype of cells.

## Methods and Materials

### Study participants

All brain specimens were derived from two longitudinal clinico-pathological cohorts: the Religious Orders Study (ROS), or the Memory and Aging Project (MAP). In both cohorts, participants did not have known dementia at the time of enrollment. The participants agreed to receive clinical evaluation each year and donate their brain after death; the two studies were designed and are run by the same group of investigators, with essentially identical ante- and post-mortem phenotypic data collection. Thus, they are designed to be analyzed jointly (9) and are referred to as “ROSMAP”. For this study, 424 participants with both snucRNAseq and whole genome sequence data were retained for analysis. At the time of death, 34% of participants were cognitively non-impaired, 26% were mildly impaired, and 40% were demented. Of the 424 participants, 68% were female.

### ROSMAP depressive symptoms

ROSMAP depressive symptoms were assessed with a modified, 10-item version of the Center for Epidemiologic Studies Depression scale (CES-D). Participants were asked whether or not they experienced each of ten symptoms much of the time in the past week (e.g. could not get going, felt depressed). The score is the total number of symptoms experience, more details can be found in https://www.radc.rush.edu/docs/var/detail.htm?category=Depression&variable=cesdsum.

In this study, we have analyzed differential gene expression based on two depressive symptom scores: the score from the last visit for each participant, and the average depressive symptom score over time calculated for each donor (averge visit times=8 (SD=±6), follow-up time frame: IQ-25=3.0yrs, median=6.2yrs, IQ-75=11.2yrs).

### Linear regression model for depressive symptom-associated gene detection

We implemented a linear regression model to identify depressive symptom-associated gene expression in 7 cell types and 55 cell subtypes (Eq. 1).

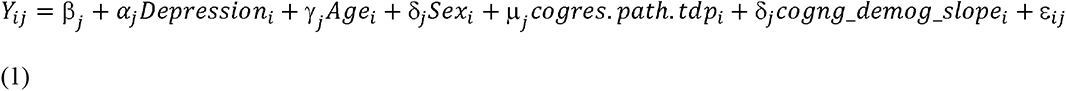

In our linear regression model, *Y_ij_* is the expression level of gene *j* in donor i, *Depression_i_* denotes the last visit or average depressive symptom score of donor *i*, *Age_i_* denotes the age in last visit or age at death of donor *i*, *Sex_i_* denotes the sex of donor *i*, *cogres. path. tdp_i_* is the residual cognition adjusted by amyloid, tau and TDP-43 pathologies of donor *i*, *cogng_demog_slope_i_* represents the random slope of global cognition of donor *i*, ε_ij_ is the error term, *α_j_, γ_j_, δ_j_, μ_j_, δ_j_*, is the correlation coefficient for each variates. The corresponding correlation and *p*-values (adjusted with BH (Benjamini & Hochberg) were then calculated for all genes; only FDR value<0.05 were considered as significant depressive symptom-associated genes.

### Whole genome sequencing

Whole genome sequencing (WGS) of ROSMAP participants was performed as described previously (10). Briefly, DNA was extracted from brain or blood samples. WGS libraries were prepared using the KAPA Hyper Library Preparation Kit and sequenced on an Illumina HiSeq X sequencer as paired end reads of 150 bp. Reads were mapped to the reference human genome GRCh37 using BWA-mem, and variants were called by GATK HaplotypeCaller. For this study, the VCF file was lifted over to GRCh38 using Picard LiftoverVcf, and only variants that passed the GATK filter (variant quality score-based sensity<99.8%) were used (11). Variant Call Format (VCF) files of GWAS are available at Synapse (https://www.synapse.org/#!Synapse:syn11724057).

### Single-nucleus RNA-seq

Single-nucleus RNA sequencing (snucRNAseq) of 424 DLPFC was performed as described previously (12). In brief, frozen specimens of DLPFC from ROSMAP participants were obtained from Rush Alzheimer’s Disease Center. Gray matter was extracted and dissociated into nuclei suspension. Single-nucleus RNA-seq libraries were constructed using Chromium Single Cell 3’ Reagent Kits version 3 (10x Genomics) following the manufacturer’s protocol and sequenced using HiSeqX and NovaSeq sequencers (Illumina). FASTQ files were processed using CellRanger (v6.0.0; 10x Genomics) with the “GRCh38-2020-A” transcriptome and the “include-introns” option. Cell calling and ambient RNA removal were performed using the CellBender software. All raw and processed data are available through the AD Knowledge Portal (https://www.synapse.org/#!Synapse:syn31512863).

### Cell type classifications

Based on the cell-type annotations of our prior work (13), we used a stepwise clustering approach to identify first the major cell types of the DLPFC and then subtypes in each cell type. In the end, we analyzed data organized into 7 major cell types, which were further subdivided into 92 cell subtypes found in the human DLPFC (more details can be found in (12)).

### Pseudo-bulk expression

For each cell type and cell subtype, pseudo-bulk UMI count matrices were constructed by extracting UMI counts of the cell (sub)type of interest, aggregating them for each participant, and normalizing them by sequencing depth. Pseudo-bulk UMI counts were normalized by using the trimmed mean of M-values (TMM) method of edgeR (14), and log_2_ of counts per million mapped reads (CPM) were computed using the voom function of limma (version 3.44.3) (15). Low expression genes (log_2_CPM<2.0) were filtered out. Batch effects were corrected using ComBat (16). Expression levels were quantile normalized. Pseudo-bulk expression of cell subtypes was quantified by the same method.L

### *cis*-eQTL mapping

*Cis-*eQTLs were identified by linear regression with 1 Mbp of the transcription start site of each measured gene (gene expression derived using log_2_CPM), as implemented in Matrix eQTL (ver. 2.3) (17), adjusting for top 3 genotype PCs, top 30 expression PCs, age, sex, post-mortem interval, study (ROS or MAP), and total number of genes detected in each participant. Multiple hypothesis correction was performed using a two-step method. Gene-wise *p*-values were computed by applying Bonferroni correction to the smallest nominal *p*-value of each gene with the number of tested SNPs for the gene. The threshold for statistical significance of eGenes was set to the false discovery rate (FDR)<5%, where FDR was computed from gene-wise *p*-values using the Benjamini-Hochberg method. Statistical significance of eSNPs were judged by nominal *p*-values, and its threshold was set to the largest nominal *p*-value of gene-SNP pair that had FDR<5%. More details can be found in our recent paper (12).

### GWAS summary statistics

We used depression GWAS summary statistics from 500,199 participants of European descent by Howard et.al (5) that did not include 23andMe participants, of these, 361,315 were from the UK Biobank and 138,884 were from the Psychiatric Genetics Consortium (18). Approximately 34% of participants had depression.

Five other diseases/traits GWAS summary statistics were used to investigate the specificity of the depression TWAS results. Including three psychiatric disorders: bipolar disorder (BD) (*N*_cases_=41,917; *N*_controls_=371,549) (19), schizophrenia (SCZ) (*N*_cases_=76,755; *N*_controls_=243,649) (20) and neuroticism (*N*=390,278) (21); body-mass-index (BMI) (*N*=∼700,000) (22) and waist-to-hip ratio adjusted for BMI (WHRadjBMI) (*N*=484,680) (23).

### Expression network analysis

Gene expression network was constructed by WGCNA (24), an R package that implements a computationally optimized procedure for weighted gene coexpression network analysis. Gene expression values were normalized and covariate-adjusted (i.e., study index, postmortem interval and total genes detected). For each gene expression module, we performed module-trait correlation analysis between module eigengene and 29 phenotypic traits (e.g. last visit/average depressive symptom score, amyloid, *APOE* genotype, braak stage). To do this, we corrected age for sex, sex for age, age and sex for the other 27 traits, and education was corrected as an extra covariate for cognition-related traits (cogng_demog_slope). To account for multiple hypotheses, FDR corrected p-value threshold (FDR<=0.05) was used to define significant trait-module relationships. Similar analysis was applied to other five cell types: astrocytes, excitatory neurons, inhibitory neurons, oligodendrocytes and OPCs. We also performed correlation analysis between each module eigengene and the frequency of cell subtypes within each major cell type.

### eQTL integrative analysis

We used pseudo-bulk RNA-seq data and genotypes from ROSMAP (424 brain subjects) to impute the *cis* genetic component of expression into the depression GWAS summary statistics (5). The complete TWAS pipeline is implemented in the FUSION (ver. Oct. 1, 2019) suite of tools (6). The details steps implemented in FUSION are as follows. First, we estimated the heritability of gene expression and stopped if not significant. We estimated using a robust version of GCTA-GREML (25), which generates heritability estimates per feature as well as the likelihood ratio test *P* value. Only features that have a heritability of *P*L<L0.05 were retained for TWAS analysis. Second, the expression weights were computed by modeling all *cis*-SNPs (±1LMb from the transcription start site) using best linear unbiased prediction, or modeling SNPs and effect sizes with Bayesian sparse linear mixed model, least absolute shrinkage and selection operator, Elastic Net and top SNPs (6, 26). A cross-validation for each of the desired models were then performed. Third, a final estimate of weights for each of the desired models was performed and the results were stored. The imputed unit is treated as a linear model of genotypes with weights based on the correlation between SNPs and expression in the training data while accounting for linkage disequilibrium (LD) among SNPs. To account for multiple hypotheses, FDR corrected *p*-value threshold (FDR ≤ 0.05) was used to define significant TWAS associations. snucRNA-sequencing from 55 cell subtypes (sample size ≥ 100) was also imputed for TWAS analysis (**Figure S1**).

SMR (ver. 1.03) (7) was used to test whether depression TWAS-significant genes (from the FUSION approach) were associated with depression via their *cis*-regulated brain transcriptomics. We used the ROSMAP genotype, our pseudo-bulk cell types eQTL results and the Howard et al. depression GWAS summary statistics to perform SMR, the conservative unadjusted *P*L<0.05 from heterogeneity in dependent instruments (HEIDI) was used to suggest that the presence of linkage likely influences the main SMR findings. Similarly, we performed FUSION, SMR and HEIDI in bipolar disorder, schizophrenia, neuroticism, BMI and WHRadjBMI.

### Colocalization analysis

The COLOC package (version 5.1.0) was applied to test the approximate Bayes factor (ABF) colocalization hypothesis, which assumes a single causal variant. Under ABF analysis, the association of a trait with a SNP is assessed by calculating the posterior probability (value from 0 to 1), with the value of 1 indicating the causal SNP. In addition, the ABF analysis has 5 hypotheses, where, PP.H0.abf indicates there is neither an eQTL nor a GWAS signal at the loci; PP.H1.abf indicates the locus is only associated with the GWAS; PP.H2.abf indicates the locus is only associated with the eQTL; PP.H3.abf indicates that both the GWAS and eQTL are associated but to a different genetic variant; PP.H4.abf indicates that the eQTL and the GWAS are associated to the same genetic variant. With the posterior probability of each SNP and aiming to find the casual variants between the GWAS and eQTL, we focused on extracting the PP.H4 value for each SNP in our study.

For depression GWAS (5), we used the reported lead SNPs of 102 loci. For each locus, we searched for the eSNPs that are within 500 Kb of the lead SNP, and listed eGenes that were paired with the eSNP. We then obtained the eGenes *cis*-eQTL output around the lead eSNP within 1 Mbp window size. In addition, we extracted GWAS summary statistics around the reported 102 lead SNP. At last, we conducted COLOC for respective pair of eGene-eQTL and eSNP-GWAS for each cell type.L

### Function annotation for cell subtype-level depression TWAS genes

The DAVID tool (27) was used to perform GO annotation. Up-/down-regulated depression TWAS gene lists were submitted to DAVID by choosing GO_FAT and KEGG pathway terms to describe the overrepresented functional terms. The threshold for overrepresented GO terms was set to FDR<0.05.

### Cross-trait LD score regression

LDSC (28) bivariate genetic correlations attribute to genome-wide SNPs (r_g_) were estimated across seven human diseases/traits from published GWASs as we mentioned above. We used LD scores from the ‘eur_w_ld_chr’ file available from https://alkesgroup.broadinstitute.org/LDSCORE, computed using 1000 Genomes Project (29) Europeans as a reference panel as previously described (30). Adjusting for the number of traits tested, the FDR *P*-value threshold (FDR<0.05) was used to define significant genetic correlations.

## Results

### Differentially expressed depressive symptom genes

The Religious Order Study (ROS) and the Memory and Aging Project (MAP) are two longitudinal studies of brain aging with annual neuropsychiatric evaluations and prospective autopsy; all participants in the two studies are non-demented at the time of enrollment. The two studies were designed and are run by a single set of investigators, and they were designed to be analyzed jointly. As a result, we refer to them as ROSMAP. The ROSMAP team has assessed the depressive symptoms of their particpants over time, we assessed the relation of each cell subtype to depressive symptom scores. Specifically, we evaluated two related outcomes: (1) the depressive symptom score at the last visit prior to death using a modified, 10-item version of the Center for Epidemiologic Studies Depression scale (CES-D), and (2) the average depressive symptom score over the participant’s time in the study (IQ-25=3.0yrs, median=6.2yrs, IQ- 75=11.2yrs) (see Methods, **Figure S2**). The latter outcome may help to better capture the particpant’s history of depression while the former outcome may best capture the state near the time of death. A linear regression model was then used to identify differentially expressed depressive symptom genes in 55 cell subtypes with pseudo-bulk RNA-seq measures (we limit these analyses to those subtypes that have >10 cells in ≥ 100 participants), adjusting for age, sex, and two manifestations of AD: slope of global cognitive function and residual cognition adjusted for AD pathologies. We adjusted for these two cognition-related traits due to their significant correlation with the last visit/average depressive symptom scores (**Supplementary Table S1**).

By using the last visit depressive symptom score and an FDR<0.05, we identified 8 depressive symptom-associated genes in astrocytes, neurons or oligodendrocytes (**Table 1**). For example, TAO Kinase 3 (*TAOK3*) (FDR=0.01) (**Figure 2**), which showed downregulated gene expression with depressive symptom in oligodendrocyte subtype 12. The protein encoded by this gene is a serine/threonine protein kinase that activates the p38/MAPK14 stress-activated MAPK cascade but inhibits the basal activity of the MAPK8/JNK cascade. A previous study found individuals suffering from bipolar disorder and schizophrenia showed that a microdeletion that affects *TAOK3* (and *PEBP1*) is present in schizophrenia patients (31). Further, a GWAS analysis suggested that *TAOK3* may contribute to neurodevelopmental disorders, at least in schizophrenia (32). Another gene, MAP kinase-activating Death Domain protein (*MADD*) showed increased expression with depressive symptom in inhibitory neuron subtype 11 (FDR=0.03). This gene plays a survival-promoting role against TNF mediated apoptosis (33), and it has been found associated with post-traumatic stress disorder (34).

**Table 1:** Results of the differentially expressed genes identified from 55 cell subtypes. Asterisks indicate genes whose cis-regulated brain mRNA levels were also associated with depression based on a transcriptome-wide association study (TWAS) of depression that integrated the depression GWAS (N=L500,199) with snucRNAseq transcriptomic and genetic data (N=424).

| Gene | Chromosome | coefficient | p-value | FDR | Cell subtype |
| --- | --- | --- | --- | --- | --- |
| **Last visit depressive symptom score** |  |  |  |  |  |
| *ADHFE1* | 8 | 0.20 | $8.43 \times 10^{-6}$ | 0.02 | Ast.5 |
| *DCUN1D2* | 13 | 0.20 | $7.85 \times 10^{-6}$ | 0.02 | Ast.5 |
| *CHUK\** | 10 | -0.14 | $2.13 \times 10^{-6}$ | 0.03 | Exc.4 |
| *AC005225.2* | 14 | -0.14 | $2.20 \times 10^{-6}$ | 0.02 | Inh.6 |
| *MADD* | 11 | 0.14 | $6.76 \times 10^{-6}$ | 0.03 | Inh.11 |
| *MEF2A* | 15 | 0.13 | $6.87 \times 10^{-6}$ | 0.03 | Oli.2 |
| *CLASP2* | 3 | 0.16 | $6.27 \times 10^{-5}$ | 0.04 | Oli.7 |
| *TAOK3\** | 12 | -0.14 | $4.29 \times 10^{-6}$ | 0.02 | Oli.12 |
| **Average depressive symptom score** |  |  |  |  |  |
| *CRADD* | 12 | -0.18 | $2.20 \times 10^{-6}$ | 0.02 | Exc.7 |
| *CCDC6\** | 10 | 0.33 | $1.18 \times 10^{-6}$ | $7.26 \times 10^{-3}$ | Exc.10 |
| *AC005225.2* | 14 | -0.17 | $4.01 \times 10^{-6}$ | 0.03 | Inh.6 |
| *C11orf58\** | 11 | 0.19 | $8.73 \times 10^{-6}$ | 0.03 | Inh.9 |
| *ZCCHC17* | 1 | 0.179 | $6.71 \times 10^{-6}$ | 0.03 | Inh.11 |
| *TBC1D5* | 3 | -0.18 | $7.84 \times 10^{-6}$ | 0.01 | Inh.12 |
| *SCAI\** | 9 | 0.19 | $4.03 \times 10^{-6}$ | 0.01 | Inh.12 |

**Figure 2.**
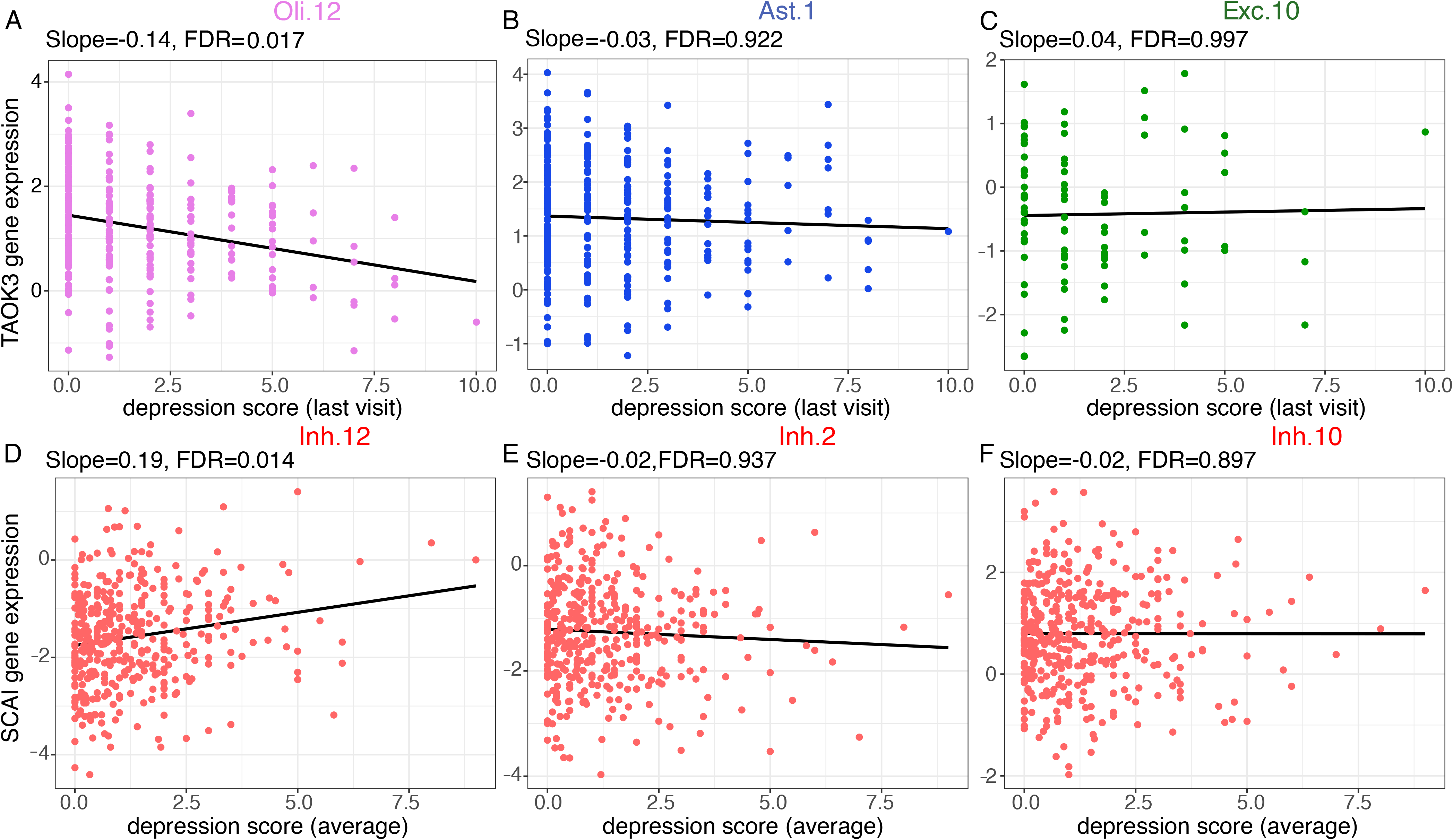
Example of depressive symptome-associated gene expression changes in cell subtypes A,B,C) Scatterplot shows depression associated *TAOK3* gene expression changes in oligodendrocyte subtype 12, astrocytes subtype 1 and excitatory neurons subtype 10, using the last visit of depressive symptom score. **D,E,F)** Scatterplot shows depression associated *SCAI* gene expression changes in inhibitory neurons subtype 12, 2 and 10, using the average depressive symptom score over time.

Another 7 depressive symptom-associated genes were identified in neurons when using the average depressive symptom score over the study period (**Table 1**), including Coiled-Coil Domain Containing 6 (*CCDC6*) (FDR=7.26×10^-3^) and Suppressor of Cancer Cell Invasion (*SCAI*) (FDR=0.01) (**Figure 2**). *CCDC6* encodes a coiled domain-containing protein, which it has been reported as a causal gene for the psychiatric and neurodegenerative diseases at both the mRNA and protein levels in bulk cortex data (35). *SCAI* was found downregulated in several human tumors, and decreased levels of *SCAI* are tightly correlated with increased invasive cell migration (36). It was also found to be modestly associated with major depressive disorders (MDD) (37).

We did not pursue pathway analyses given the small number of associated genes, which are not adequate to support such analyses. Thus, while we are likely still underpowered for a differential gene expression analysis, we identified a small number of intriguing candidates, including two neuronal genes that have been implicated in prior studies of bulk cortical data. To be thorough, we also assembled a list of 109 statistically robust reported associations from bulk cortical RNA data in prior studies (38–40), and we evaluated them for altered expression in our data: we detected no significant differential expression. However, this is not surprising, since depression is a polygenic trait, arising from the influence of multiple loci with small individual effects, and our sample size remains modest.

To complement the single gene analyses, we repeated our analyses after reducing the dimensionality of the data: using WGCNA, we established modules of co-expressed genes in each cell type and derived an eigengene measure for each module (**Figure S3A-C, Supplementary Table S2**). However, these analyses returned no association to either the last measure or the average measure of depressive symptom (**Figure S3B**), suggesting that, as with GWAS, we will need substantially larger studies to be properly powered for identifying alterations in gene expression relative to depression. Finally, we also assessed the frequency of 107 cell subtypes that we defined in these data (12) in relation to the two outcomes, but we did not find any significant associations either. Thus, while direct association analyses in our moderately sized study for depression yielded only a small number of results, this data resource is well-powered for other analyses – such as expression quantitative trait locus (eQTL) mapping - that can be leveraged to powerfully interrogate the genetic architecture of the disease.

### A Transcriptome-Wide Association Study (TWAS) of depression

We integrated our snucRNAseq-based eQTL results (12) with the latest depression GWAS (5) results to perform a TWAS of depression using the FUSION pipeline (6): the expression level of a subset of genes can be inferred from single nucleotide polymorphism (SNP) data, and these genetic instruments can be deployed into any relevant collection of genome-wide genotype data. Using over 1.6 million human brain transcriptomes derived from individual nuclei extracted from the DLPFC of 424 older individuals (**Figure S4**) of European descent from the ROSMAP cohorts (41), we have previously mapped *cis*-eQTLs in 7 major cell types, and 55 cell subtypes which have data from more than 100 participants (12). Genes that had significant single nucleotide polymorphism (SNP)-based heritability (p<0.05) were retained in the TWAS analysis (**Supplementary S3, Figure S5**). Depression GWAS summary statistics were from 500,199 participants of European descent, who were not 23andMe, Inc. participants. TWAS identified the greatest number of associated genes in excitatory neurons: 99 genes displayed association of inferred gene expression with depression (FDR<0.05) (**Figure 3A** **and Supplementary Table S4**). Associations in other cell types include: 68 genes in inhibitory neurons, 67 genes in astrocytes, 51 genes in oligodendrocytes, 33 genes in oligodendrocyte progenitor cells (OPCs), 28 genes in microglia, and 2 genes in endothelial cells. We note that our dataset had relatively few endothelial cell nuclei in each participant, limiting our power to map *cis*-eQTLs in that cell type; in general, the less frequent cell types such as microglia also had fewer genes whose expression we could infer (12).

**Figure 3.**
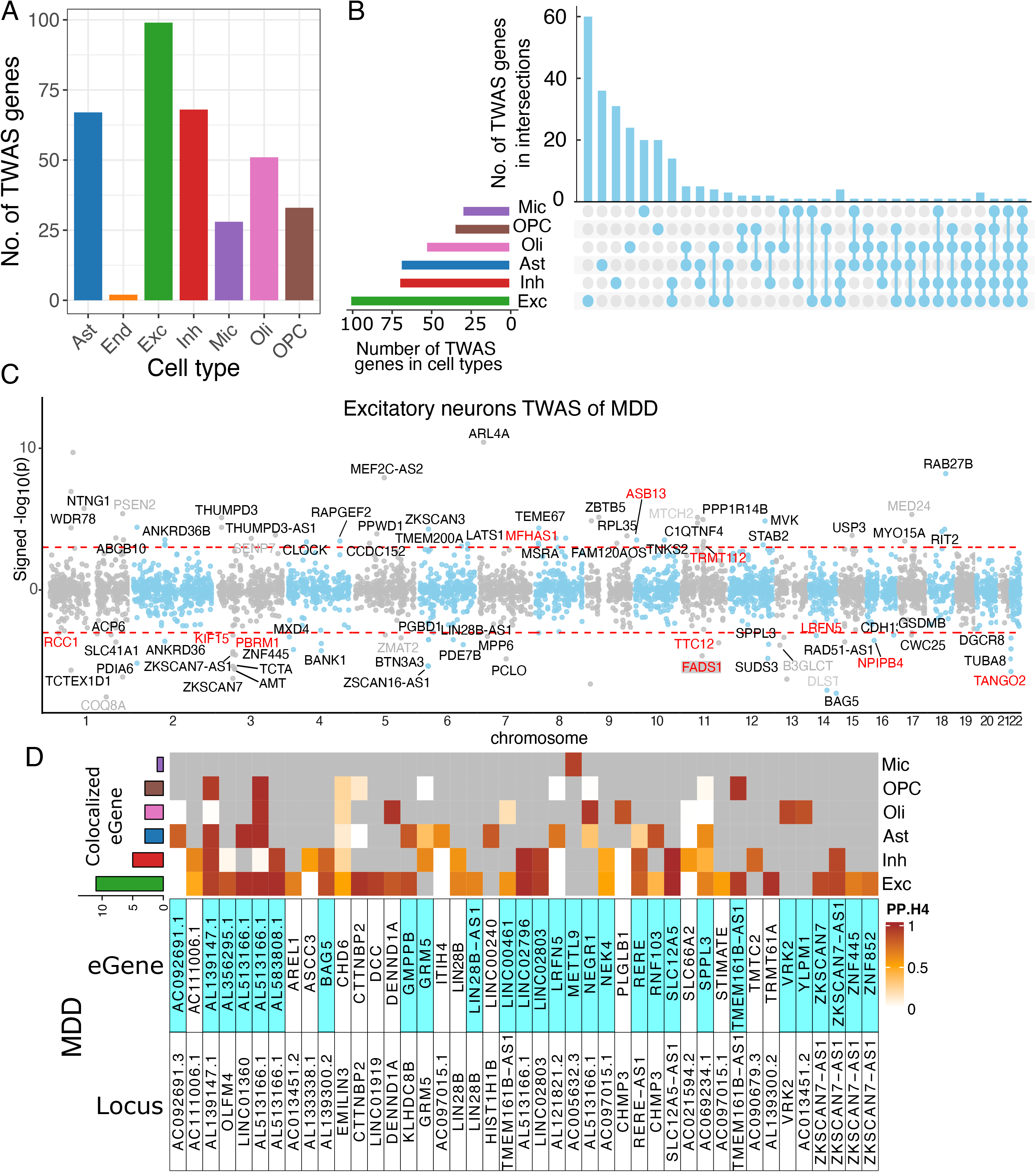
Depression genes identified from pseudo-bulk RNA-seq. **A)** Number of depression TWAS genes identified from 7 cell types. **B)** Number of depression TWAS genes that were unique to or shared between 7 cell types. **C)** Statistical significance and effect directions of excitatory neurons TWAS of depression. Each dot represents a gene. Positive and negative y coordinates show that transcript abundance was associated with increased and decreased risk of depression, respectively. Novel and known candidates for depression risk protein-coding genes are colored black and gray, respectively. Genes in red color are TWAS genes filtered with SMR p<0.05 & HEIDI p>0.05. Red dashed lines show genes with TWAS FDR<0.05. **D)** Heatmap reports the posterior probabilities of the H4 hypothesis (PP.H4) of the COLOC method, which assumes GWAS and eQTL share a single causal SNP. Rows report overlap for individual genes and SNP pair; columns report PP.H4 score in each of our cell types. The color of each cell is based on the code found to the right of each panel; the darker color denotes higher confidence that the same variant influences susceptibility and gene expression in that cell type. Grey cells indicate that the gene was not an eQTL target in that cell type. Top bar chart shows the number of colocalized eGenes with high confidence (PP.H4>0.5) in each cell type. Cells highlighted with blue color indicates that the same gene was also detected by cell type-level TWAS.

To further evaluate whether gene expression mediates the association between genetic variants and depression for each of the genes identified in one of the cell types in the FUSION analyses, we performed Summary-data-based Mendelian Randomization (SMR) (7). Results from SMR demonstrated that 67/99 genes in excitatory neurons, 36/68 genes in inhibitory neurons, 34/67 genes in astrocytes, 30/51 genes in oligodendrocytes, 16/33 in OPC, 15/28 in microglia reached a nominally significant level (p<0.05). Next, we used the heterogeneity independent instrument (HEIDI) test to distinguish pleiotropy/causality from linkage for genes in each of the 7 cell types. HEIDI results suggested that 11/99 genes in excitatory neurons, 6/68 genes in inhibitory neurons, 4/67 genes in astrocytes, 4/51 genes in oligodendrocytes, 2/33 genes in OPC, 3/18 genes in microglia were consistent with either pleiotropy or causality (p>0.05) (**Supplementary Table S5**). In **Figure 3C**, we illustrate a Manhattan plot of the 99 depression TWAS genes identified in excitatory neurons, highlighting the results of novel candidates as well as SMR and HEIDI results.

Next, we conducted colocalization analysis (COLOC) to assess whether altered gene expression may be the mechanism for a particular risk allele or haplotype. Using a genome-wide association study of major depressive disorder (MDD) and our list of eGenes, we find evidence of colocalization (PP.H4>0.5) for 43 eGenes among the 102 MDD loci that we interrogated (**Figure 3D, Supplementary Table S6**). As expected, excitatory and inhibitory neurons harbor the most implicated target genes. We note that while many loci have unambiguous cell type-specific effects (i.e., *TMTC2* or *ZKSCAN7*), *AL139147*.1 has an effect shared by four cell types (Inh, Exc, Ast, Oli and OPC). In addition, the COLOC analysis is helpful in supporting the FUSION analysis: 65% (28/43) of TWAS genes also showed colocalized effects in MDD. Taken together, our results from FUSION, SMR and HEIDI, and COLOC anlaysis suggested that there is an enrichment of depression genes in neurons and astrocytes.

We also applied FUSION, SMR and HEIDI on the 55 cell subtypes of single-nucleus RNA-seq for identifying depression genes (**Supplementary Table S7-S9**). Consistent with results at the cell type level, we identified the greatest number of depression TWAS genes in excitatory neurons, especially subtype 4, 12 and 3 (**Supplementary Table S7**). Among the 1,068 depression TWAS genes identified from 55 cell subtypes, 167 of them were found shared with 252 genes in 7 cell types (**Supplementary Table S10**). The high but not complete overlap of depression TWAS genes (66.3%) identified from cell types and cell subtypes indicates that using cell subtype-level snucRNAseq data can provide additional gene signals whose expression is altered in specific cell subtypes.

Overall, our results showed that the effect directions of genetically regulated expression on depression risk are largely consistent across cell subtypes within the same cell type (**Figure 4A**). For example, the increased gene expression of *SLC12A5* was found associated with increased depression risk across cell subtypes of excitatory neurons and inhibitory neurons. A previous study integrating depression GWAS with bulk brain and blood eQTL data found *SLC12A5* expression showed evidence of a genetically predicted effect on depression and neuroticism (42), so we refined this robust association by highliting the relevant cell type. By contrast, *NEGR1* in excitatory neuron subtype 3 (Exc.3) and 4 (Exc.4) showed opposite effects for genetically regulated expression of depression risk: in Exc.3, the increased gene expression of *NEGR1* is associated with increased depression risk, while in subtype 4, the increased gene expression of *NEGR1* associated with decreased depression risk (**Figure 4A**). *NEGR1* is a cell adhesion molecule expression in neurons, and its expression in primary hippocampal neurons was found to be significantly reduced in antidepressant-treated rats (43). Similar patterns of opposite directions of effect can be seen with *ZFYVE21* and *B3GLCT* in excitatory neuron subtypes, and *TMEM161B-AS1* in inhibitory neuron subtypes (**Figure 4A**). This is intriguing, suggesting that gene expression related to functional genetic variation can be differentially modulated among different subtypes of neurons.

**Figure 4.**
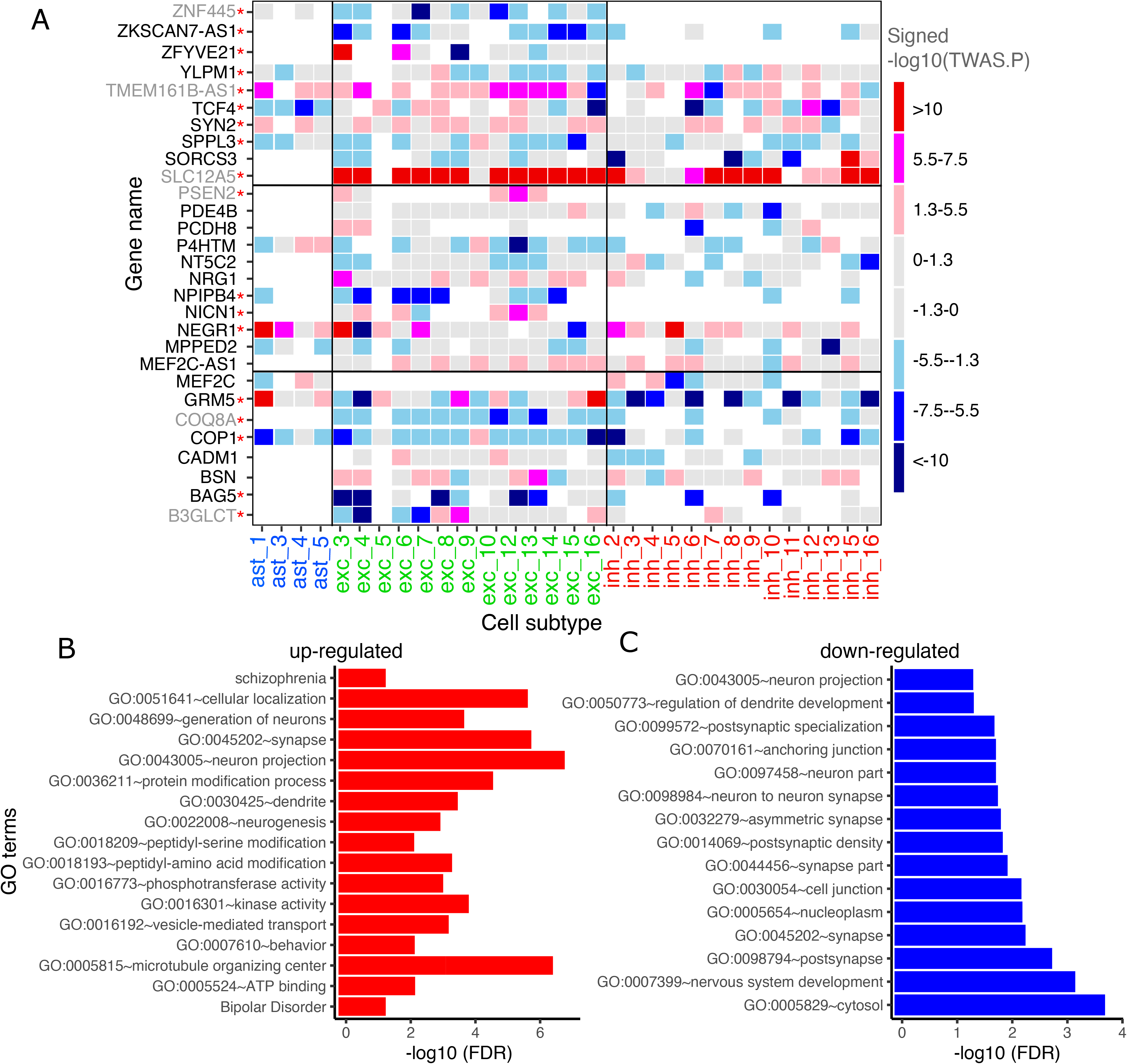
Cell subtype-level TWAS of depression. **A)** Subtype-level pseudo-bulk expression and GWAS summary statistics of Howard et al., 2019 were used as input. Non-gray elements show 30 significant TWAS genes (FDR<0.05), and their colors represent effect directions and p-values. In row names, novel and known candidates for depression risk genes are colored black and gray, respectively. Red asterisk indicates that the same gene was also detected by cell-type level TWAS. Grey color represents the genes which are not significant in TWAS analysis (TWAS FDR>0.05). White color denotes genes that did not detected in the corresponding cell subtype. Genes were ordered alphabetically from bottom to top. **B)** GO terms and KEGG pathways enriched in the up-regulated genes associated with depression. **C)** GO terms and KEGG pathways enriched in the down-regulated genes associated with depression.

In addition, we inspected the functional similarity and difference among genes associated with depression by using the DAVID tool (27). Depresion TWAS genes were divided into 491 with up- and 449 with down-regulation with respect to the effect of depression risk allele, and genes showed inconsistent effect directions across cell subtypes were excluded for this analysis. GO annotation and pathway analysis demonstrated differential functional enrichment, among up- regulated depression TWAS genes (**Figure 4B**): they were characterized by being involved in neuronal projection (false discovery rate (FDR=1.01×10^-7^), synapse function (FDR=1.09×10^-6^), protein serine/threonine kinase activity (FDR=2.18×10^-3^), behavior (FDR=4.39×10^-3^), or bipolar disorder and schizophrenia (FDR=0.03). On the other hand, down-regulated depression TWAS genes were found to be enriched in fewer functions (**Figure 4C**), for example cytosol (FDR=1.51×10^-4^), nervous system development (FDR=5.21×10^-4^), nucleoplasma (FDR=4.77×10^-3^) and anchoring junction (FDR=0.01) (a full list of functional annotations is provided in **Supplementary Table S11**). These results suggest that depression variants where the risk allele increases gene expression may be more likely to influence neuronal function at the synapse while the loci containing alleles that diminish expression will need further evaluation.

### Identifying novel depression genes

To extract novel depression genes identified from our analysis, we compiled a list of 176 unique genes reported from previous studies (18, 44, 45), which all used a TWAS approach in bulk RNAseq data. Comparing these known depression TWAS genes with our FUSION, SMR and HEIDI results, we found in 7 cell types, 34/252 (13.5%) of depression TWAS genes (hypergeometric test p<1.00×10^-16^), and 14/43 (32.6%) of depression COLOC genes were common with the known candidates, while this number reduced to 5/28 (17.9%) when using the SMR and HEIDI results (hypergeometric test p=0.32). When it comes to the 55 cell subtypes, we found 67/1,068 (6.3%) of the depression TWAS genes (hypergeometric test p<1.00×10^-16^), and 9/51 (17.6%) of TWAS-significant SMR and HEIDI genes (hypergeometric test p<3.20×10^-03^) are shared with the known gene list from bulk RNA studies. Thus, combining cell types and cell subtypes results, we identified 59 unique novel depression genes based on FUSION, SMR and HEIDI analysis (**Supplementary Table S12**). This represents an important step torwards enabling a cell type-specific downstream dissection of genes implicated in susceptibility to depression.

### Cell type similarity and specificity of depression TWAS genes

We next investigated the specificity of depression genes by displaying the extent to which they are shared among the different cell types (**Figure 3B**). The results showed a cell type-specific pattern, for example, 60/99 (60.6%) depression genes in excitatory neurons were specific to this cell type, and only 14 of the 99 genes were found to be shared with inhibitory neurons. This result can be explained by (i) a target gene that is expressed only in one cell type, (ii) a target gene that is expressed in multiple cell types but where the genetic variant affects only one cell type. A similar trend was observed in the results of the cell subtype analysis (**Figure S6**).

### Trait similarity and specificity of the depression TWAS genes

To understand the trait specificity of the depression TWAS results, we performed a TWAS for neuroticism, bipolar disorder (BD), schizophrenia (SCZ), body-mass-index (BMI) and waist-to-hip ratio (WHR) adjusted BMI (WHRadjBMI) (**Figure 5A**). These traits were chosen because they have a range of estimated genetic correlation with depression: 0.69 for neuroticism, 0.45 for BD, 0.32 for SCZ, 0.11 for BMI and WHRadjBMI (**Figure 5B**). We expected that traits with evidence of higher genetic correlation would have more TWAS results in common. Next, we performed TWAS analysis on GWAS summary statistics for each trait (see Methods) and pseudo-bulk RNA-seq in seven cell types. Combining 7 cell types together, the TWAS of neuroticism identified 521 genes, the TWAS of BD identified 574 unique genes, the TWAS of SCZ identified 1,318 unique genes, the TWAS of BMI identified 57 unique genes and the TWAS of WHRadjBMI identified 3,033 genes (**Supplementary Table S13**).

**Figure 5.**
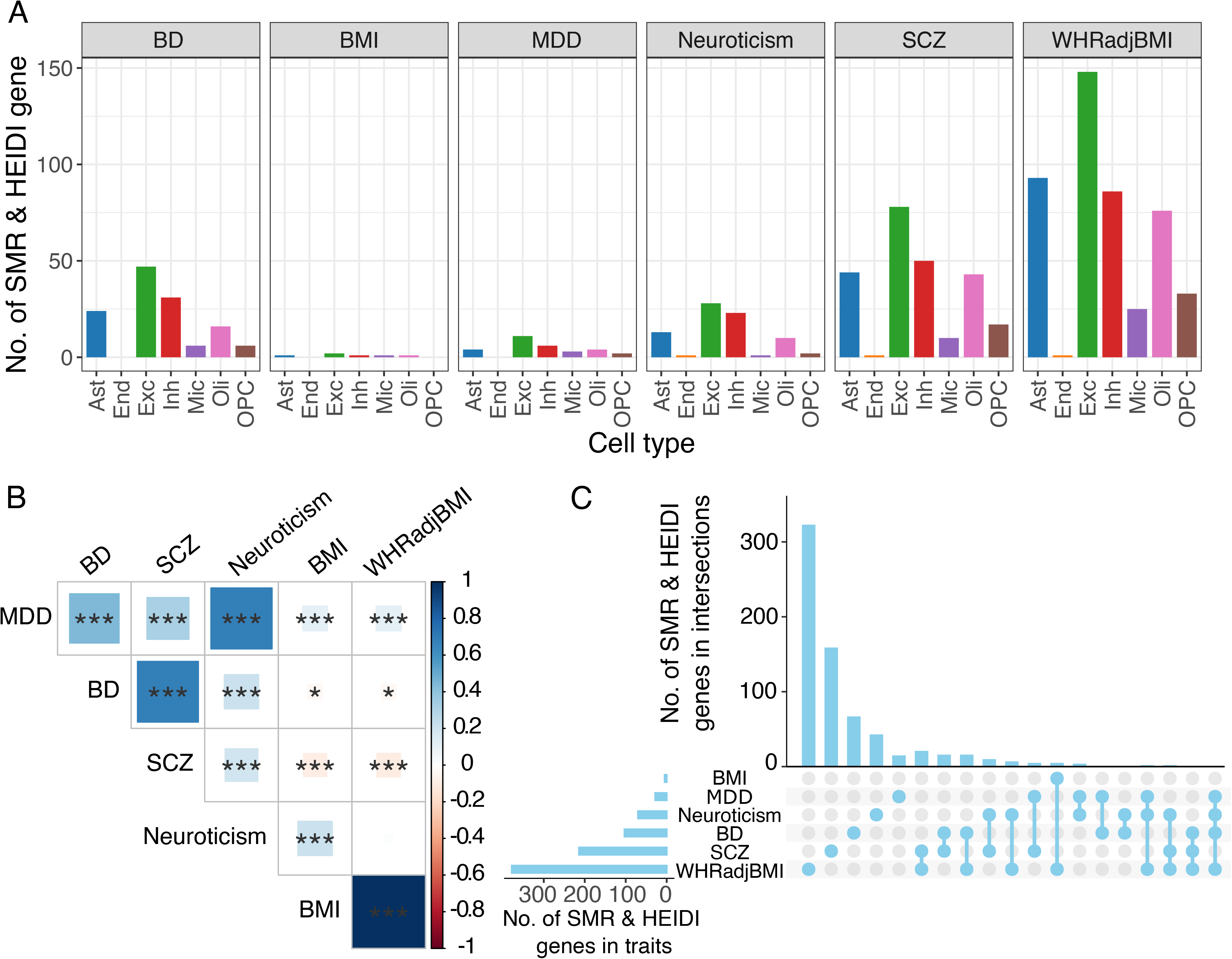
Trait specificity of depression TWAS genes filtered with SMR p<0.05 & HEIDI p>0.05. **A)** Number of TWAS, SMR and HEIDI genes identified from 7 cell types of pseudo-bulk RNA- seq associated with six traits (bipolar disorder (BD), body mass index (BMI), major depressive disorder (MDD), neuroticism, schizophrenia (SCZ), and waist-to-hip ratio adjusted for BMI (WHRadjBMI)). **B)** Genetic correlation estimated between depression and other diseases/traits. The areas of the squares represent the absolute value of corresponding genetic correlations. After FDR correction for 36 tests at 5% significance level, genetic correlations estimates that are significantly different from zero are marked with a asterisk (*0.01<FDR<=0.05; **0.001<FDR<=0.01; ***FDR<=0.001). The blue color denotes a positive genetic correlation, the red color represents a negative genetic correlation. **C)** Number of TWAS, SMR and HEIDI genes that were unique to or shared between other diseases/traits.

In addition, we applied SMR and HEIDI on the TWAS-significant genes to remove genes with an SMR p<0.05, a HEIDI p>0.05 or in cases where a HEIDI *P* value was unable to be determined. These tests allow us to focus on genes with evidence that their genetically regulated gene expression mediates their association with the trait of interest and to remove genes likely to be the result of linkage disequilibrium. After considering findings from FUSION, SMR and HEIDI, we identified 70 genes in neuroticism, 103 genes in BD, 214 genes in SCZ, 5 genes in BMI and 378 genes in WHRadjBMI that likely contribute to these traits by modulating the brain transcriptome (**Supplementary Tables S13**). Unsurprisingly, 7 of 70 (10.00%) of the neuroticism genes, 2 of 103 (1.94%) of the BD genes, 5 of 214 (2.34%) of the SCZ genes were also identified by the depression TWAS, which reflects the genetic correlation among these traits.

By contrast, 0 of 5 (0.00%) of the BMI genes and 3 of 378 (0.79%) of the WHRadjBMI genes overlapped with the 28 depression TWAS-significant genes.

Additionally, our results showed a trait-specific pattern for our analyses of inferred gene expression, for example, 15/28 (53.6%) of depression genes, 159/214(74.3%) of SCZ genes, 323/378 (85.4%) of WHRadjBMI genes were found specific to itself (**Figure 5C**).

## Discussion

In this study, we prioritized genes expressed in specific neocortical cell types and subtypes that contribute to the pathogenesis of depression to accelerate the prioritization of new therapeutic targets. We analyzed data from the DLPFC, a region of the brain that is an important hub for mood-related circuits. We used multiple complementary approaches, including differential gene expression analysis as well as methods integrating our data with GWAS results using FUSION, SMR and HEIDI, and COLOC. Overall, leveraging the known genetic architecture of depression, we identified 71 causal genes in depression and have identified those cell types in which the functional consequences are occurring. 59 of these genes are novel compared to previous studies, for example, *ANKRD36*, *PBRM1* and *FAM120AOS*. We found depression TWAS genes showed a cell type-specific pattern, with the greatest enrichment being in neurons and astrocytes, in comparison to Alzheimer’s disease where we have reported an excess of microglial genes (46). Depression TWAS genes were found to be more often shared with TWAS genes for traits that are more genetically correlated (e.g., neuroticism) with depression when compared with traits with lower genetic correlation (e.g., BMI).

Five genes (*MADD*, *TAOK3*, *SCAI*, *C11orf58* and *CHUK*) were both identified by the TWAS and differential expression analysis. *MADD* plays a survival-promoting role against TNF mediated apoptosis (33), it has been found associated with post-traumatic stress disorder (34). A previous study found individuals suffering from bipolar disorder and schizophrenia showed that a microdeletion that affects *TAOK3* (and *PEBP1*) is present in schizophrenia patients (31), and a GWAS analysis suggested that *TAOK3* alone may contribute to neurodevelopmental disorders, at least in schizophrenia (32). *SCAI* was found downregulated in several human tumors, and decreased levels of *SCAI* are tightly correlated with increased invasive cell migration (36). It was also found modest associated with major depressive disorders (37). *C11orf58* was found associated with bipolar disorder in whole brain expression (47), and *CHUK* plays a key role in the negative feedback of NF-kappa-B canonical signaling to limit inflammatory gene activation, where the defective expression of NF- kappa-B has been proposed to play a role in the development of depression (48).

In recent years, the target cell types in depression pathophysiology expanded from excitatory neurons to include inhibitory interneurons (49) and non-neuronal cells (38, 50, 51). Previous studies have reported that the greatest depression-associated differential gene expression occurred in deep layer excitatory neurons and immature oligodendrocyte precursor cells (OPCs), which contributed almost half (47%) of all changes in gene expression (52). The density and form of cell abnormalities (in astrocytes, microglia, or oligodendrocytes) play an important role in psychiatric disorders, including BD, depression, and SCZ (53). Here, we found depression, SCZ and BD TWAS genes showed the greatest enrichment in excitatory neurons, followed by inhibitory neurons, astrocytes, oligodendrocytes, OPC and microglia. In terms of cell subtypes, excitatory neurons 3,4 and 12 showed the greatest enrichment of depression genes, as well as inhibitory neurons 2, 6 and 15. Our results further confirmed the pre-eminent role of neurons in depression but also highlights that at least some of the functional consequences of risk variants are likely to be widely distributed among cortical cell subtypes, and our analyses provide evidence that certain risk variants and gene expression may have opposite functional effects on depression in different neuron subtypes. These data support the idea that there is functional heterogeneity among neurons, and that many additional cell subtype resolved analyses will be needed to fully map the functional consequences of depression susceptibility loci.

Despite of an excess of common TWAS genes between depression and other psychiatric disorders (neuroticism, SCZ and BP) when compared to less genetically correlated traits such as BMI and WHRadjBMI. We found >53% of TWAS genes were only identified in depression and 74.3% of TWAS genes were specific to SCZ, indicating genetic regulated gene expression distinctions between psychiatric disorders. These findings could help with development of treatment biomarkers targeting each specific psychiatric disorder.

In summary, this study illustrates the utility of large snucRNAseq data to uncover both genes whose expression is altered in specific cell subtypes in the context of depression and to enhance the interpretation of well-powered GWAS so that we can prioritize specific susceptibility genes for further analysis.

## Data Availability

ROSMAP resources can be requested at https://www.radc.rush.edu.

## Acknowledgements and Disclosures

The study was supported by NIH grants P30AG10161, P30AG72975, R01AG15819, R01AG17917, U01AG46152, U01AG61356, RF1 AG057473, U01 AG072572. ROSMAP resources can be requested at https://www.radc.rush.edu. The content is solely the responsibility of the authors and does not necessarily represent the official views of the National Institutes of Health.

LZ and PLD conceived and designed the study; LZ performed the research; MF, ZG, CCW and GSG provided data and code contributed to the analyses; HUK, DAB, PAB, NH and VM participated in the discussion and interpretation of the results; LZ and PLD wrote the paper; LZ created the figures which were edited by PLD; All the authors reviewed and revised the paper.

## Notes

### Competing Interest Statement

The authors have declared no competing interest.

### Author Declarations

All of the studies were approved by the institutional review board of Rush University, Columbia University, and Partners Healthcare/Broad Institute. Informed consent was received from all participants or their representatives.

## Reference

1. Friedrich MJ. Depression Is the Leading Cause of Disability Around the World. JAMA. 2017;317(15):1517.

2. Disease GBD, Injury I, Prevalence C. Global, regional, and national incidence, prevalence, and years lived with disability for 310 diseases and injuries, 1990-2015: a systematic analysis for the Global Burden of Disease Study 2015. Lancet. 2016;388(10053):1545–602.

3. Edwards SL, Beesley J, French JD, Dunning AM. Beyond GWASs: illuminating the dark road from association to function. Am J Hum Genet. 2013;93(5):779–97.

4. Gamazon ER, Segre AV, van de Bunt M, Wen X, Xi HS, Hormozdiari F, et al. Using an atlas of gene regulation across 44 human tissues to inform complex disease- and trait-associated variation. Nat Genet. 2018;50(7):956–67.

5. Howard DM, Adams MJ, Clarke TK, Hafferty JD, Gibson J, Shirali M, et al. Genome-wide meta-analysis of depression identifies 102 independent variants and highlights the importance of the prefrontal brain regions. Nat Neurosci. 2019;22(3):343–52.

6. Gusev A, Ko A, Shi H, Bhatia G, Chung W, Penninx BW, et al. Integrative approaches for large-scale transcriptome-wide association studies. Nat Genet. 2016;48(3):245–52.

7. Zhu Z, Zhang F, Hu H, Bakshi A, Robinson MR, Powell JE, et al. Integration of summary data from GWAS and eQTL studies predicts complex trait gene targets. Nat Genet. 2016;48(5):481–7.

8. Giambartolomei C, Vukcevic D, Schadt EE, Franke L, Hingorani AD, Wallace C, et al. Bayesian test for colocalisation between pairs of genetic association studies using summary statistics. PLoS Genet. 2014;10(5):e1004383.

9. Bennett DA, Schneider JA, Buchman AS, Barnes LL, Boyle PA, Wilson RS. Overview and findings from the rush Memory and Aging Project. Curr Alzheimer Res. 2012;9(6):646–63.

10. De Jager PL, Ma Y, McCabe C, Xu J, Vardarajan BN, Felsky D, et al. A multi-omic atlas of the human frontal cortex for aging and Alzheimer’s disease research. Sci Data. 2018;5:180142.

11. Van der Auwera GA, Carneiro MO, Hartl C, Poplin R, Del Angel G, Levy-Moonshine A, et al. From FastQ data to high confidence variant calls: the Genome Analysis Toolkit best practices pipeline. Curr Protoc Bioinformatics. 2013;43:11 0 1- 0 33.

12. Fujita M, Gao Z, Zeng L, McCabe C, White CC, Ng B, et al. Cell-subtype specific effects of genetic variation in the aging and Alzheimer cortex. bioRxiv. 2022:2022.11.07.515446.

13. Anael Cain MT, Cristin McCabe, Idan Hekselman, Charles C. White, Gilad Green, Orit Rozenblatt-Rosen, Feng Zhang, Esti Yeger-Lotem, David A. Bennett, Hyun-Sik Yang, Aviv Regev, Vilas Menon, Naomi Habib, Philip L. De Jager. Multi-cellular communities are perturbed in the aging human brain and with Alzheimer’s disease. bioRxiv. 2020.

14. Robinson MD, McCarthy DJ, Smyth GK. edgeR: a Bioconductor package for differential expression analysis of digital gene expression data. Bioinformatics. 2010;26(1):139–40.

15. Ritchie ME, Phipson B, Wu D, Hu Y, Law CW, Shi W, et al. limma powers differential expression analyses for RNA-sequencing and microarray studies. Nucleic Acids Res. 2015;43(7):e47.

16. Johnson WE, Li C, Rabinovic A. Adjusting batch effects in microarray expression data using empirical Bayes methods. Biostatistics. 2007;8(1):118–27.

17. Shabalin AA. Matrix eQTL: ultra fast eQTL analysis via large matrix operations. Bioinformatics. 2012;28(10):1353–8.

18. Wray NR, Ripke S, Mattheisen M, Trzaskowski M, Byrne EM, Abdellaoui A, et al. Genome-wide association analyses identify 44 risk variants and refine the genetic architecture of major depression. Nat Genet. 2018;50(5):668–81.

19. Mullins N, Forstner AJ, O’Connell KS, Coombes B, Coleman JRI, Qiao Z, et al. Genome-wide association study of more than 40,000 bipolar disorder cases provides new insights into the underlying biology. Nat Genet. 2021;53(6):817–29.

20. Trubetskoy V, Pardinas AF, Qi T, Panagiotaropoulou G, Awasthi S, Bigdeli TB, et al. Mapping genomic loci implicates genes and synaptic biology in schizophrenia. Nature. 2022;604(7906):502–8.

21. Nagel M, Jansen PR, Stringer S, Watanabe K, de Leeuw CA, Bryois J, et al. Meta-analysis of genome-wide association studies for neuroticism in 449,484 individuals identifies novel genetic loci and pathways. Nat Genet. 2018;50(7):920–7.

22. Yengo L, Sidorenko J, Kemper KE, Zheng Z, Wood AR, Weedon MN, et al. Meta-analysis of genome-wide association studies for height and body mass index in approximately 700000 individuals of European ancestry. Hum Mol Genet. 2018;27(20):3641–9.

23. Pulit SL, Stoneman C, Morris AP, Wood AR, Glastonbury CA, Tyrrell J, et al. Meta-analysis of genome-wide association studies for body fat distribution in 694 649 individuals of European ancestry. Hum Mol Genet. 2019;28(1):166–74.

24. Langfelder P, Horvath S. WGCNA: an R package for weighted correlation network analysis. BMC Bioinformatics. 2008;9:559.

25. Yang J, Lee SH, Goddard ME, Visscher PM. GCTA: a tool for genome-wide complex trait analysis. Am J Hum Genet. 2011;88(1):76–82.

26. Gusev A, Mancuso N, Won H, Kousi M, Finucane HK, Reshef Y, et al. Transcriptome-wide association study of schizophrenia and chromatin activity yields mechanistic disease insights. Nat Genet. 2018;50(4):538–48.

27. Sherman BT, Hao M, Qiu J, Jiao X, Baseler MW, Lane HC, et al. DAVID: a web server for functional enrichment analysis and functional annotation of gene lists (2021 update). Nucleic Acids Res. 2022.

28. Bulik-Sullivan B, Finucane HK, Anttila V, Gusev A, Day FR, Loh PR, et al. An atlas of genetic correlations across human diseases and traits. Nat Genet. 2015;47(11):1236–41.

29. Genomes Project C, Auton A, Brooks LD, Durbin RM, Garrison EP, Kang HM, et al. A global reference for human genetic variation. Nature. 2015;526(7571):68–74.

30. Finucane HK, Bulik-Sullivan B, Gusev A, Trynka G, Reshef Y, Loh PR, et al. Partitioning heritability by functional annotation using genome-wide association summary statistics. Nat Genet. 2015;47(11):1228–35.

31. Malhotra D, McCarthy S, Michaelson JJ, Vacic V, Burdick KE, Yoon S, et al. High frequencies of de novo CNVs in bipolar disorder and schizophrenia. Neuron. 2011;72(6):951–63.

32. Gilman SR, Chang J, Xu B, Bawa TS, Gogos JA, Karayiorgou M, et al. Diverse types of genetic variation converge on functional gene networks involved in schizophrenia. Nat Neurosci. 2012;15(12):1723–8.

33. Kurada B, Li LC, Mulherkar N, Subramanian M, Prasad KV, Prabhakar BS. MADD, a splice variant of IG20, is indispensable for MAPK activation and protection against apoptosis upon tumor necrosis factor-alpha treatment. J Biol Chem. 2009;284(20):13533–41.

34. Zhang Z, Meng P, Zhang H, Jia Y, Wen Y, Zhang J, et al. Brain Proteome-Wide Association Study Identifies Candidate Genes that Regulate Protein Abundance Associated with Post-Traumatic Stress Disorder. Genes (Basel). 2022;13(8).

35. Wingo TS, Liu Y, Gerasimov ES, Vattathil SM, Wynne ME, Liu J, et al. Shared mechanisms across the major psychiatric and neurodegenerative diseases. Nat Commun. 2022;13(1):4314.

36. Brandt DT, Baarlink C, Kitzing TM, Kremmer E, Ivaska J, Nollau P, et al. SCAI acts as a suppressor of cancer cell invasion through the transcriptional control of beta1-integrin. Nat Cell Biol. 2009;11(5):557–68.

37. Mostafavi S, Battle A, Zhu X, Potash JB, Weissman MM, Shi J, et al. Type I interferon signaling genes in recurrent major depression: increased expression detected by whole-blood RNA sequencing. Mol Psychiatry. 2014;19(12):1267–74.

38. Pantazatos SP, Huang YY, Rosoklija GB, Dwork AJ, Arango V, Mann JJ. Whole-transcriptome brain expression and exon-usage profiling in major depression and suicide: evidence for altered glial, endothelial and ATPase activity. Mol Psychiatry. 2017;22(5):760–73.

39. Darby MM, Yolken RH, Sabunciyan S. Consistently altered expression of gene sets in postmortem brains of individuals with major psychiatric disorders. Transl Psychiatry. 2016;6(9):e890.

40. Mahajan GJ, Vallender EJ, Garrett MR, Challagundla L, Overholser JC, Jurjus G, et al. Altered neuro-inflammatory gene expression in hippocampus in major depressive disorder. Prog Neuropsychopharmacol Biol Psychiatry. 2018;82:177–86.

41. Bennett DA, Buchman AS, Boyle PA, Barnes LL, Wilson RS, Schneider JA. Religious Orders Study and Rush Memory and Aging Project. J Alzheimers Dis. 2018;64(s1):S161–S89.

42. Korologou-Linden R, Leyden GM, Relton CL, Richmond RC, Richardson TG. Multi-omics analyses of cognitive traits and psychiatric disorders highlights brain-dependent mechanisms. Hum Mol Genet. 2021.

43. Carboni L, Pischedda F, Piccoli G, Lauria M, Musazzi L, Popoli M, et al. Depression-Associated Gene Negr1-Fgfr2 Pathway Is Altered by Antidepressant Treatment. Cells. 2020;9(8).

44. Wingo TS, Liu Y, Gerasimov ES, Gockley J, Logsdon BA, Duong DM, et al. Brain proteome-wide association study implicates novel proteins in depression pathogenesis. Nat Neurosci. 2021;24(6):810–7.

45. Li X, Su X, Liu J, Li H, Li M, andMe Research T, et al. Transcriptome-wide association study identifies new susceptibility genes and pathways for depression. Transl Psychiatry. 2021;11(1):306.

46. Hansen DV, Hanson JE, Sheng M. Microglia in Alzheimer’s disease. J Cell Biol. 2018;217(2):459–72.

47. McCarthy MJ, Liang S, Spadoni AD, Kelsoe JR, Simmons AN. Whole brain expression of bipolar disorder associated genes: structural and genetic analyses. PLoS One. 2014;9(6):e100204.

48. Napetschnig J, Wu H. Molecular basis of NF-kappaB signaling. Annu Rev Biophys. 2013;42:443–68.

49. Northoff G, Sibille E. Why are cortical GABA neurons relevant to internal focus in depression? A cross-level model linking cellular, biochemical and neural network findings. Mol Psychiatry. 2014;19(9):966–77.

50. Nagy C, Suderman M, Yang J, Szyf M, Mechawar N, Ernst C, et al. Astrocytic abnormalities and global DNA methylation patterns in depression and suicide. Mol Psychiatry. 2015;20(3):320–8.

51. Edgar N, Sibille E. A putative functional role for oligodendrocytes in mood regulation. Transl Psychiatry. 2012;2:e109.

52. Nagy C, Maitra M, Tanti A, Suderman M, Theroux JF, Davoli MA, et al. Single-nucleus transcriptomics of the prefrontal cortex in major depressive disorder implicates oligodendrocyte precursor cells and excitatory neurons. Nat Neurosci. 2020;23(6):771–81.

53. Writing Committee for the Attention-Deficit/Hyperactivity D, Autism Spectrum D, Bipolar D, Major Depressive D, Obsessive-Compulsive D, and Schizophrenia EWG, et al. Virtual Histology of Cortical Thickness and Shared Neurobiology in 6 Psychiatric Disorders. JAMA Psychiatry. 2021;78(1):47–63.

