## Supplementary figures for "A single-nucleus transcriptome-wide association study implicates novel genes in depression pathogenesis": ZengL_supplementary_figures.docx

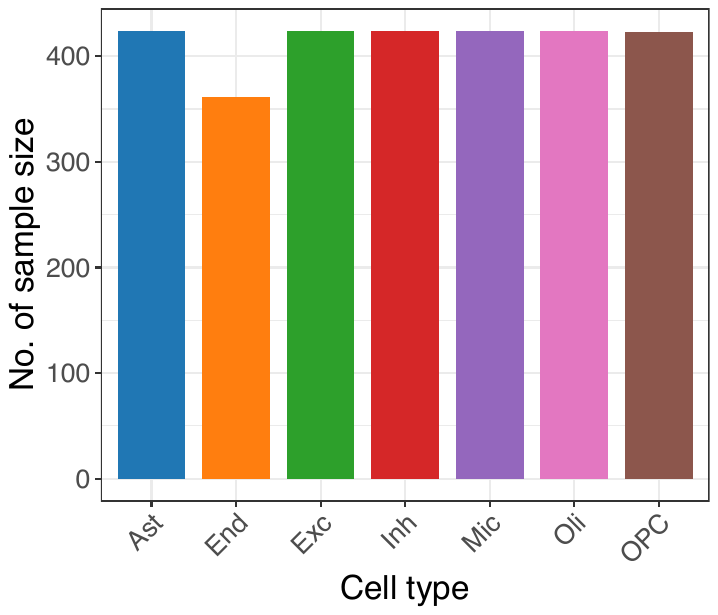

A

B

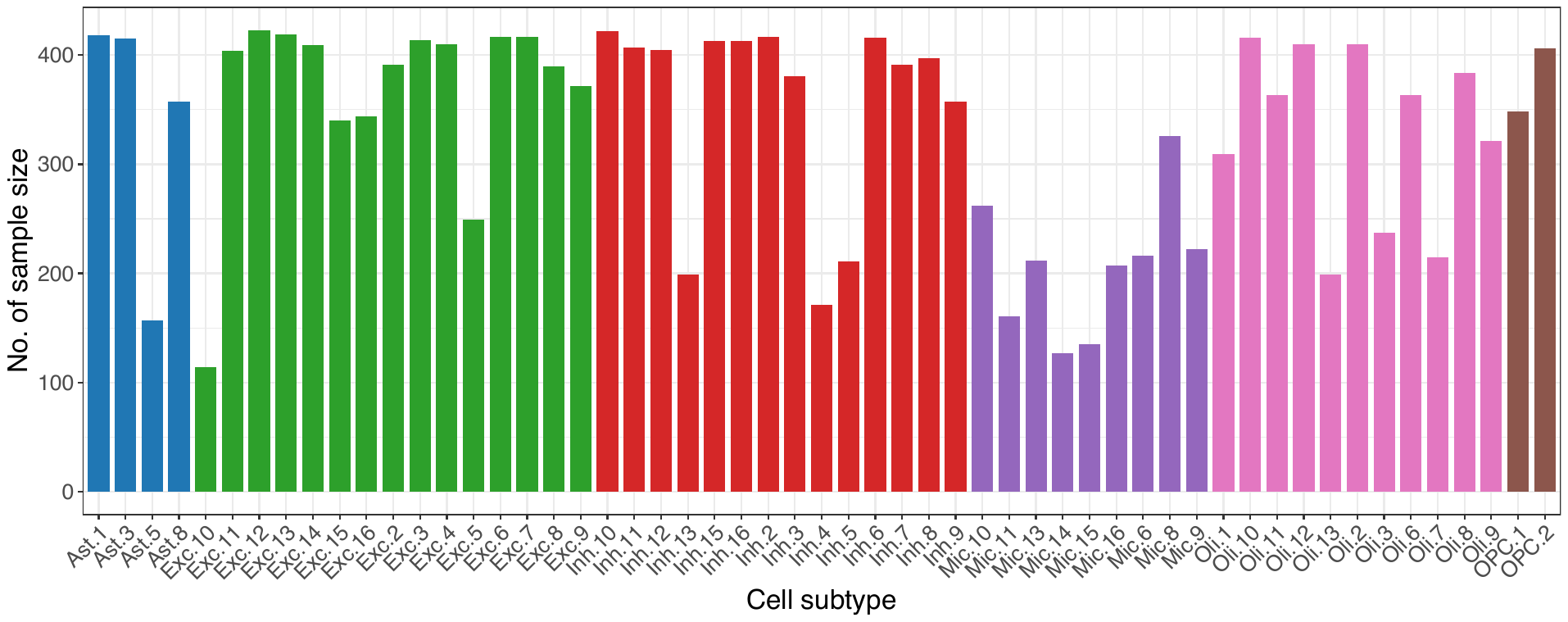

**Figure S1. The distribution of sample size used in this study**

A) Number of sample size in 7 cell types.

B) Number of sample size in 55 cell subtypes.

A

B

C

D

**Figure S2. Characteristics of the ROSMAP depressive symptom score used in this study**

**A)** Distribution of the last visit depressive symptom scores of each donor.

**B)** Distribution of the last visit depressive symptom scores *vs.* the donor’s age at their last visit.

**C)** Distribution of the average depressive symptom scores over time calculated from each donor.

**D)** Distribution of the average depressive symptom scores *vs.* the donor’s age at death.

Depression symptoms were assessed with a modified, 10-item version of the Center for Epidemiologic Studies Depression scale (CES-D). Details can be found at: https://www.radc.rush.edu/docs/var/detail.htm?category=Depression&variable=cesdsum

A

B

C

**Figure S3. Cell type gene co-expression analysis**

**A)** The number of gene co-expression modules identified by WGCNA, and the number of genes in each module.

**B)** Results of WGCNA showing module-trait relationships of 29 phenotypic traits (e.g. last visit/average depressive symptom score, amyloid, *APOE* genotype and AD-related phenotypes) in excitatory neurons. Correlations of traits with modules are shown by a color-scale with green showing negative correlation and red, positive correlation. The correlation coefficients are shown at the top of each row. The corresponding FDR for each module is displayed at the bottom of each row within parentheses.

**C)** Results of WGCNA showing module-fraction relationships of frequency of 16 cell subtypes in excitatory neurons. Correlations of traits with modules are shown by a color-scale with green showing negative correlation and red, positive correlation. The correlation coefficients are shown at the top of each row. The corresponding FDR for each module is displayed at the bottom of each row within parentheses.

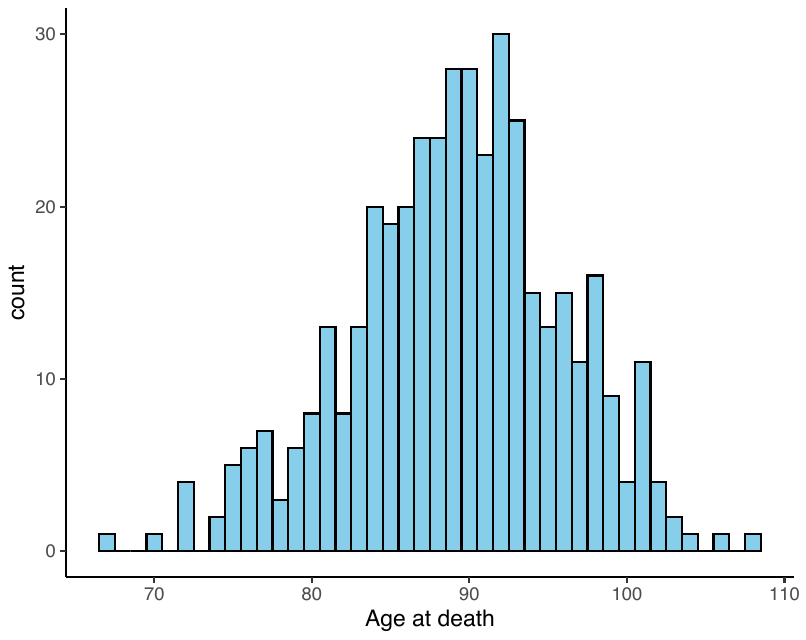

**Figure S4. The age distribution of 424 ROSMAP donors**

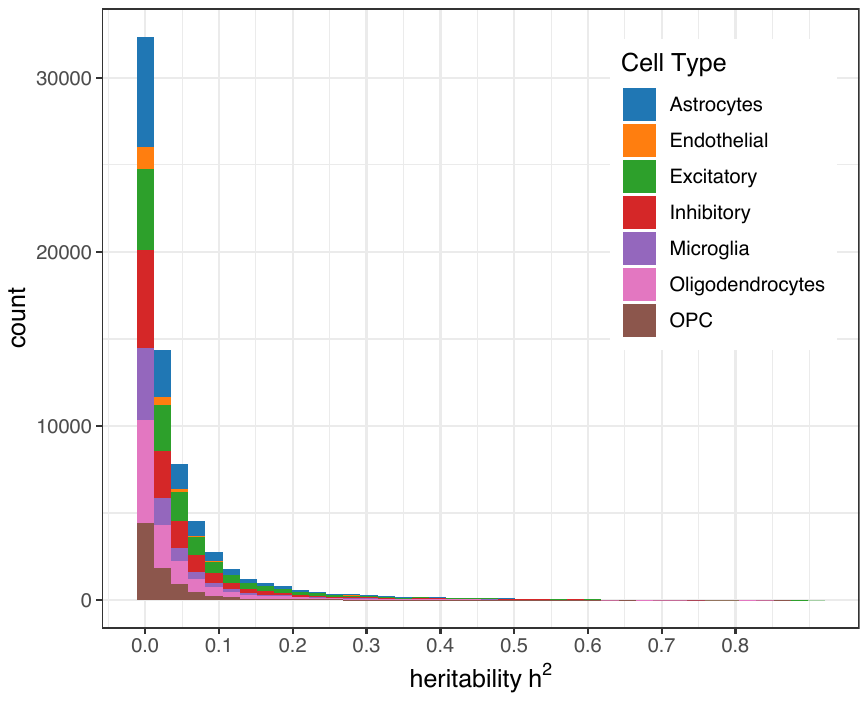

**Figure S5. Number of heritable genes identified in 7 cell types**

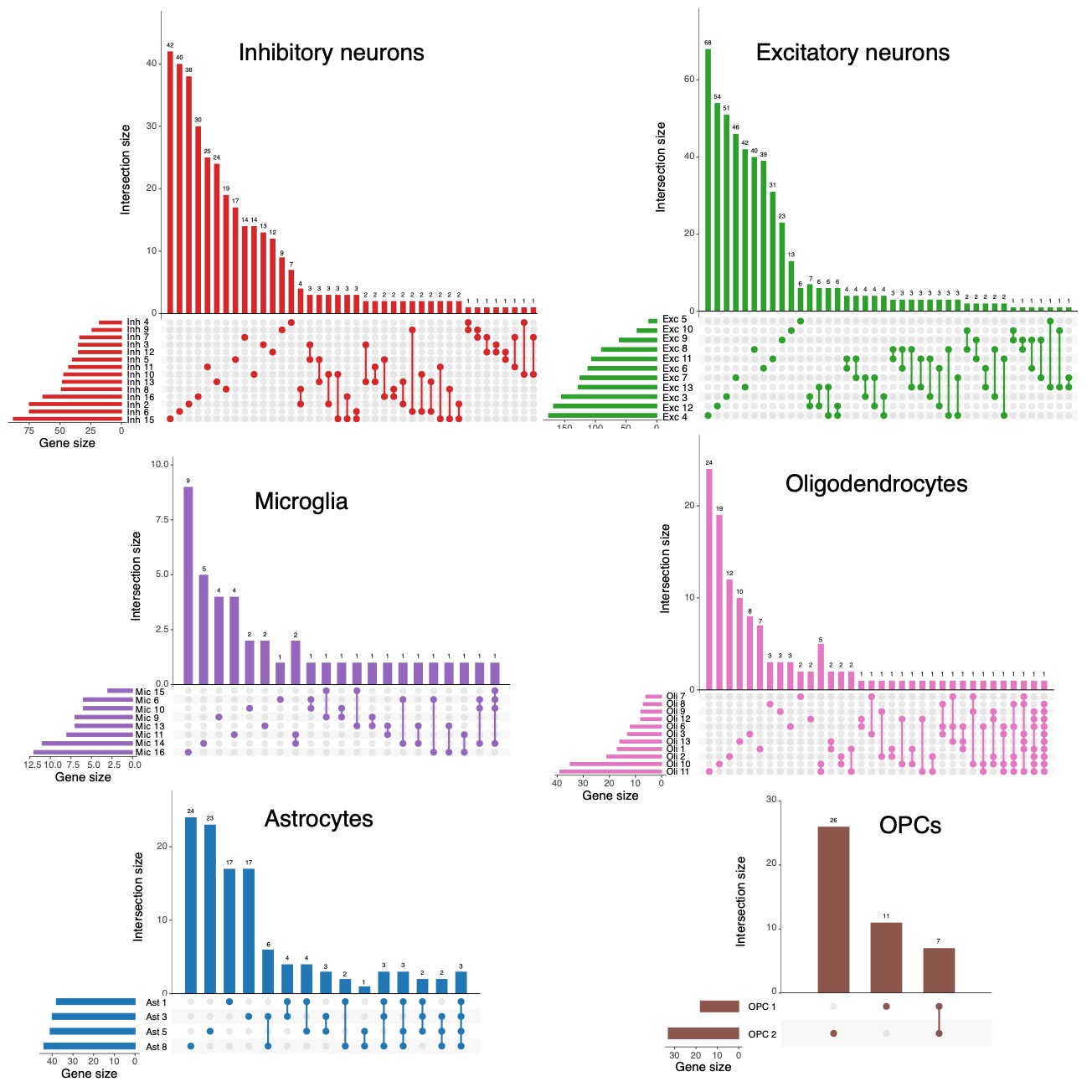

**Figure S6. Cell-subtype specificity of depression TWAS genes.**

Number of TWAS depression genes that were unique to or shared between cell subtypes.
